# Changing incidence of invasive pneumococcal disease in infants less than 90 days of age before and after introduction of the 13-valent Pneumococcal Conjugate Vaccine in Blantyre, Malawi: a 14-year hospital based surveillance study

**DOI:** 10.1101/2021.08.18.21262215

**Authors:** Marianne Koenraads, Todd D. Swarthout, Naor Bar-Zeev, Comfort Brown, Jacquline Msefula, Brigitte Denis, Queen Dube, Stephen B. Gordon, Robert S. Heyderman, Melissa Gladstone, Neil French

**Affiliations:** Department of Women and Children’s Health, Institute of Translational Medicine, University of Liverpool, Liverpool, UK; Malawi-Liverpool-Wellcome Trust Clinical Research Programme, Kamuzu University of Health Sciences, Blantyre, Malawi; NIHR Global Health Research Unit on Mucosal Pathogens, Research Department of Infection, Division of Infection and Immunity, University College London, London, United Kingdom; Institute of Infection Veterinary and Ecological Science, University of Liverpool, Liverpool, United Kingdom; International Vaccine Access Center, Department of International Health, Bloomberg School of Public Health, Johns Hopkins University, Baltimore, MD, United States; Department of Paediatrics, Kamuzu University of Health Sciences, Blantyre, Malawi; Liverpool School of Tropical Medicine, Liverpool, United Kingdom

**Keywords:** Invasive Pneumococcal Disease, Infant, Pneumococcal Conjugate Vaccine

## Abstract

**Background:** Invasive pneumococcal disease (IPD) in young infants is uncommon but associated with high morbidity and mortality. Accurate data on the burden of IPD in young infants in low-income countries are lacking. We examined the burden of IPD in infants aged <90 days in Blantyre, Malawi over a 14 year period and evaluated the impact of the 12 November 2011 introduction of the 13-valent pneumococcal conjugate vaccine (PCV13) on vaccine-serotype IPD (VT-IPD) in this population.

**Methods:** We conducted laboratory-based prospective IPD surveillance in infants aged <90 days admitted to Queen Elizabeth Central Hospital (QECH) in Blantyre between 2005 and 2018, including 7 years pre- and 7 years post-PCV13 introduction. IPD was defined as *Streptococcus pneumoniae* identified by culture from blood or cerebrospinal fluid. Serotypes were determined by multiplex PCR and latex agglutination testing.

**Results:** We identified 130 cases of culture-confirmed IPD in infants <90 days old between 2005-2018. Total IPD incidence was declining prior to PCV13 introduction. The mean incidence of IPD was significantly lower in the post-PCV era. Serotypes 5 (27.8%) and 1(15.6%), were most prevalent. Even after PCV13 introduction, VT-IPD remained dominant with serotype 5 accounting for 17.4% and serotype 1 for 13% of cases in young infants.

**Conclusion:** Vaccine serotypes were the main cause of IPD in neonates and young infants, both before and after PCV13 introduction. Further strategies need to be considered to protect this vulnerable population, including maternal or neonatal immunization and implementation of an alternative PCV schedule with a booster dose.

**Summary:** The incidence of invasive pneumococcal disease in infants in Blantyre, Malawi has declined over the past decade and more significantly after introduction of the pneumococcal conjugate vaccine. Vaccine serotypes have remained the main cause of disease in this population.

## Background

*Streptococcus pneumoniae* is a major cause of serious bacterial infections including pneumonia, sepsis and meningitis in young children. Globally, there were an estimated 294,000 pneumococcal deaths in HIV-uninfected children aged 1-59 months in 2015, with the majority occurring in sub-Saharan Africa and Asia (1). *Streptococcus pneumoniae* is considered an uncommon but well-recognized cause of invasive bacterial disease in neonates and young infants and has been associated with high morbidity and mortality, with a case fatality rate of up to 14.3% (2-6). Globally, the burden of neonatal IPD has been estimated at 36.0 per 100,000 live births in the pre-PCV period (7). However, accurate data on the burden of IPD in neonates and young infants are lacking, especially in low income countries.

In Malawi, the under-5 child mortality rate has reduced by two-thirds between 1990 and 2015, with the country therefore achieving Millennium Development Goal (MDG) 4 (8). However, neonatal mortality remains high at 22/1000 live births (9). Severe bacterial infections contribute significantly as a leading cause of death in the neonatal population (10). A previous study examining the aetiology of neonatal sepsis in Blantyre, Malawi from 1996-2001 showed that *Streptococcus pneumoniae* was responsible for 10% of neonatal sepsis cases and 23% of neonatal meningitis cases (11). Supported by GAVI, the Vaccine Alliance, the 13-valent pneumococcal conjugate vaccine (PCV13) was introduced in Malawi in November 2011 as part of the national expanded program of immunisation (EPI) with at a 3+0 schedule (at 6, 10 and 14 weeks of age). A recent study by Swarthout *et al* (12) has shown that in Malawi, despite a reduction in VT carriage after PCV13 introduction, there remains a high persistent residual carriage with all PCV13 serotypes isolated despite high vaccine uptake. In this study, we evaluated the burden of IPD in infants aged <90 days admitted to QECH in Blantyre, Malawi in the pre- and post-PCV13 period.

## Methods

### Study setting

Malawi is a small landlocked country in southern sub-Saharan Africa. It has few natural resources and is consistently ranked by the World Bank in the lowest income category (13). QECH is a large government funded district and referral hospital in Blantyre and has about 25,000 paediatric admissions a year. QECH provides free medical care to the 1.3 million urban, peri-urban and rural residents of Blantyre District.

### Case Definitions

We analysed all archived bacterial isolates from blood and cerebrospinal fluid (CSF) of infants less than 90 days old admitted to Queen Elizabeth Central Hospital (QECH) in Blantyre, southern Malawi between 1 January 2005 and 31 December 2018. IPD cases were defined as isolation of *Streptococcus pneumoniae* from a normally sterile site (i.e. blood or CSF). We defined those with a positive CSF culture as “meningitis” and those with only a positive blood culture as “bacteraemia”. There were several cases with had a positive CSF culture as well as blood culture, these were classed as “meningitis”. However although it is standard practice to take a CSF sample in all young infants with suspected sepsis, this may not have been done in all individuals. Cases in infants ≤7 days old were defined as “early-onset” disease and cases in infants 8-89 days old as “late-onset” disease. Demographic data included age and sex. We were unable to collect data on clinical outcomes or on immunization status for vaccine age-eligible infants.

### Case ascertainment and laboratory confirmation

In accordance with longstanding clinical guidelines, all young infants presenting to QECH with fever (axillary temperature >37·5°C) or clinical suspicion of sepsis or meningitis undergo blood cultures and, where appropriate, lumbar puncture. We have been conducting sentinel surveillance for laboratory-confirmed bloodstream infection and meningitis (including IPD) in all age groups at QECH since 1998, as previously described (14-16).

Specimens were processed at the co-located Malawi-Liverpool-Wellcome Clinical Research Programme laboratory, using BD BACTEC™ (Becton Dickinson, Franklin Lakes, NJ, USA). Those positive by BACTEC™ were Gram stained and cocci further assessed using the catalase test. All pneumococcal isolates were archived on Microbank™ beads (ProLab Diagnostics) at -80°C. For subsequent serotyping, archived pneumococcal isolates were plated on gentamicin-sheep blood agar (SBG; 7% sheep blood, 5µL gentamicin/mL) and incubated overnight at 37°C in 5% CO_2_. *S. pneumoniae* growth was confirmed by colony morphology and optochin disc (Oxoid, Basingstoke, UK) susceptibility. The bile solubility test was used on isolates with no or intermediate (zone diameter <14mm) optochin susceptibility. A single colony of confirmed pneumococcus was selected and grown on a new SBG plate as before. Growth from this second plate was used for serotyping by latex agglutination (ImmuLex™ 7-10-13-valent Pneumotest; Statens Serum Institute, Denmark). The ImmuLex™ kit allows for differential identification of each PCV13 VT but not for differential identification of non-VT (NVT) serotypes; NVT and non-typeable isolates are, therefore, reported as NVT. Nucleic acid amplification-based serotyping was performed on samples collected between 1 January 2009 and 31 December 2013, using the ‘Triplex sequential real-time PCR-serotyping Africa’ protocol of the Centers for Disease Control and Prevention.^24^ A random selection of serotyped isolates was sent for confirmatory serotyping by Quellung reaction at the regional pneumococcal reference laboratory at the National Institute for Communicable Disease in Johannesburg, South Africa. Since 13 August 2011, serotyping occurs in real time with specimen processing. Isolates collected before 13 August 2011 were retrospectively serotyped. Demographic information was collected at the time of sampling. Clinical data were not prospectively collected.

### Statistical analysis

We used descriptive statistics for demographic and clinical characteristics. Incidence rates were calculated by using the annual number of VT, NVT and total (VT+NVT) IPD cases in infants <90 days old multiplied by 100,000 and this divided by the annual age-specific population estimates for Blantyre. Population estimates were obtained from the 1998 and 2018 National Population Projections by Malawi’s National Statistical Office (NSO) (17, 18). We used linear interpolation of the intercensal period to estimate the year-by-year population estimates of children <1 year old and estimated the population of infants < 90 days old by taking a proportion (3/12) of this. Confidence intervals were estimated using the modified Wald method. Incidence rate ratios (IRRs) for invasive pneumococcal disease were calculated over the study duration by log-binomial regression using years (365·25 days) between study start and PCV13 introduction (2011) and from PCV13 introduction to the end of the study period, coded as a single time variable.

## Results

We identified a total 130 cases of confirmed IPD in infants <90 days old over the study’s 14-year period, 1 January 2005 – 31 December 2018. The median age at hospital presentation was 30 days. Among the 130 infant cases, 93 (71.5%) presented with meningitis and 36 (27.7%) presented with early-onset disease (0-7 days old); among which 21 (58.3%) were meningitis and 15 (41.7%) bacteraemia. Among the 94 (72.3%) infants with late-onset disease (8-89 days old), 72 (76.6%) were meningitis and 22 (23.4%) bacteraemia. A total of 104 cases of IPD occurred before the November 2011 introduction of PCV13. Twenty-five (24.0%) of these were early-onset and 79 (76.0%) late-onset disease. Among the 26 IPD cases in the post-PCV era, 11 (42.3%) were early-onset and 15 (57.7%) late-onset disease (Table 1).

**Table 1:**
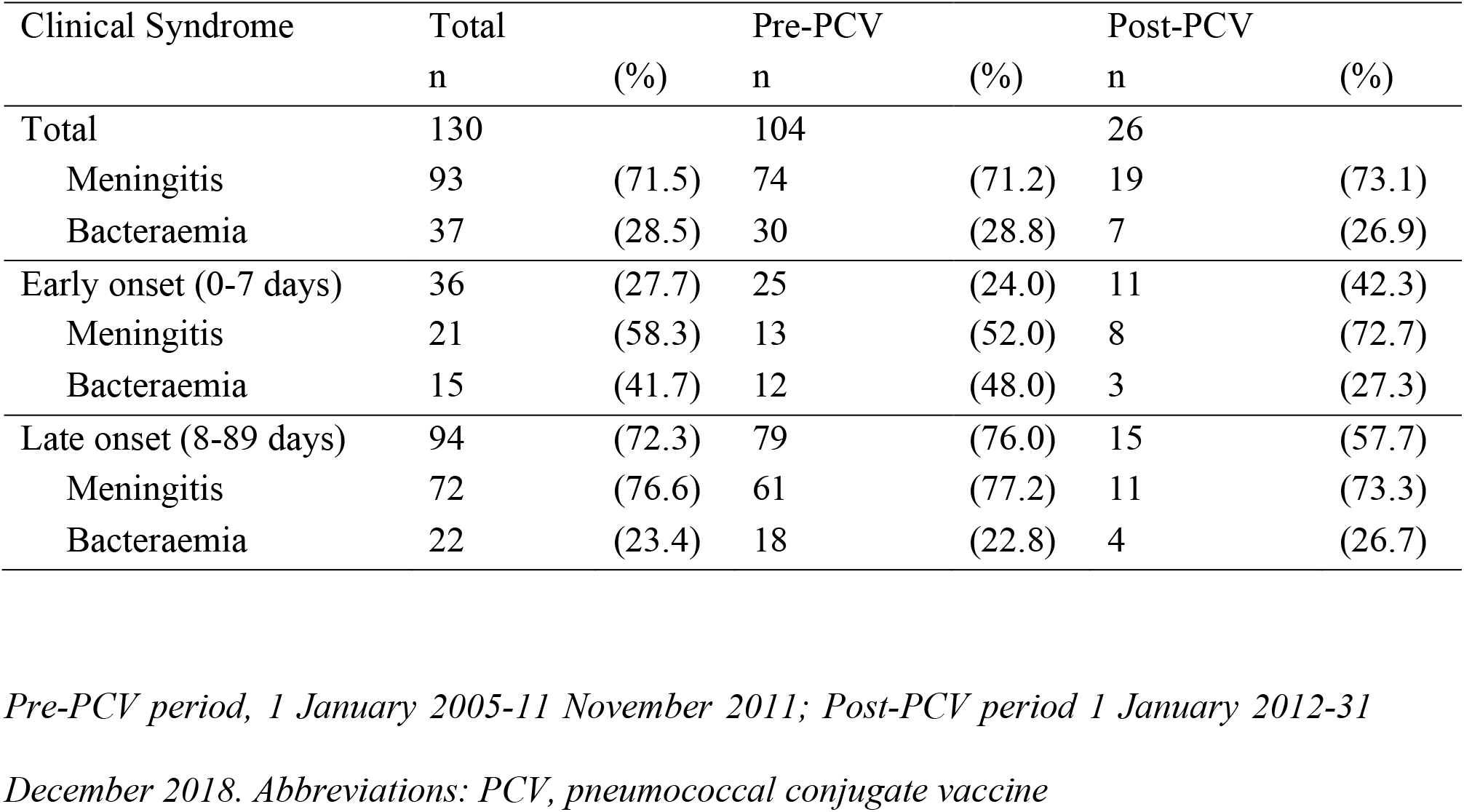
Early vs late onset disease in the Pre- and Post-PCV periods.

### Serotype distribution

Among the total 130 cases, we were able to recover and serotype 90 (69.2%) samples. Analysis of isolates that were and were not recoverable showed no statistically significant difference in age, gender, or sample type (data not shown). Over the duration of the study period, vaccine serotypes 5 (27.8%) and 1 (15.6%) were the most common in this population, with NVT-IPD accounting for 32.2% of the total recovered samples. In the pre-PCV13 period, 70.1% of IPD cases were caused by VT and 29.9% by NVT pneumococcus. In the post-PCV13 period 56.5% were VT and 43.5% NVT. The most frequent serotypes in the post-PCV13 period remained vaccine serotypes 5 (17.4%) and 1 (13.0%) (Table 2).

**Table 2:** Serotype distribution in infants <90 days in the pre-PCV and post-PCV period.

| Serotype | Total IPD |  | Pre-PCV |  | Post-PCV |  |
| --- | --- | --- | --- | --- | --- | --- |
|  | n | (%) | n | (%) | n | (%) |
| 1 | 14 | (15.6) | 11 | (16.4) | 3 | (13) |
| 3 | 2 | (2.2) | 2 | (3.0) | 0 | (0) |
| 4 | 1 | (1.1) | 1 | (1.5) | 0 | (0) |
| 5 | 25 | (27.8) | 21 | (31.3) | 4 | (17.4) |
| 6A/B | 4 | (4.4) | 3 | (4.5) | 1 | (4.3) |
| 7F | 5 | (5.6) | 4 | (6) | 1 | (4.3) |
| 9V | 0 | (0) | 0 | (0) | 0 | (0) |
| 14 | 2 | (2.2) | 0 | (0) | 2 | (8.7) |
| 18C | 3 | (3.3) | 3 | (4.5) | 0 | (0) |
| 19A | 0 | (0) | 0 | (0) | 0 | (0) |
| 19F | 0 | (0) | 0 | (0) | 0 | (0) |
| 23F | 4 | (4.4) | 2 | (3.0) | 2 | (8.7) |
| VT Total | 61 | (67.8) | 47 | (70.1) | 13 | (56.5) |
| NVT Total | 29 | (32.2) | 20 | (29.9) | 10 | (43.5) |
| Unrecovered | 40 | -- | 37 | -- | 3 | -- |
| TOTAL | 130 | -- | 104 | -- | 26 | -- |

*Pre-PCV period, 1 January 2005-11 November 2011; Post-PCV period 1 January 2012-31*
*December 2018. Abbreviations: PCV, pneumococcal conjugate vaccine; VT, vaccine serotype;*
*NVT, non-vaccine serotype.*

### IPD incidence

We estimated the annual IPD incidence rates per 100,000 infants <90 days old in Blantyre over the 14-year period. IPD incidence was already declining prior to the 2011 introduction of PCV13. After introduction of PCV13 there was a further decline in IPD cases in young infants (Figure 1; Supplementary Table 1).

**Figure 1.**
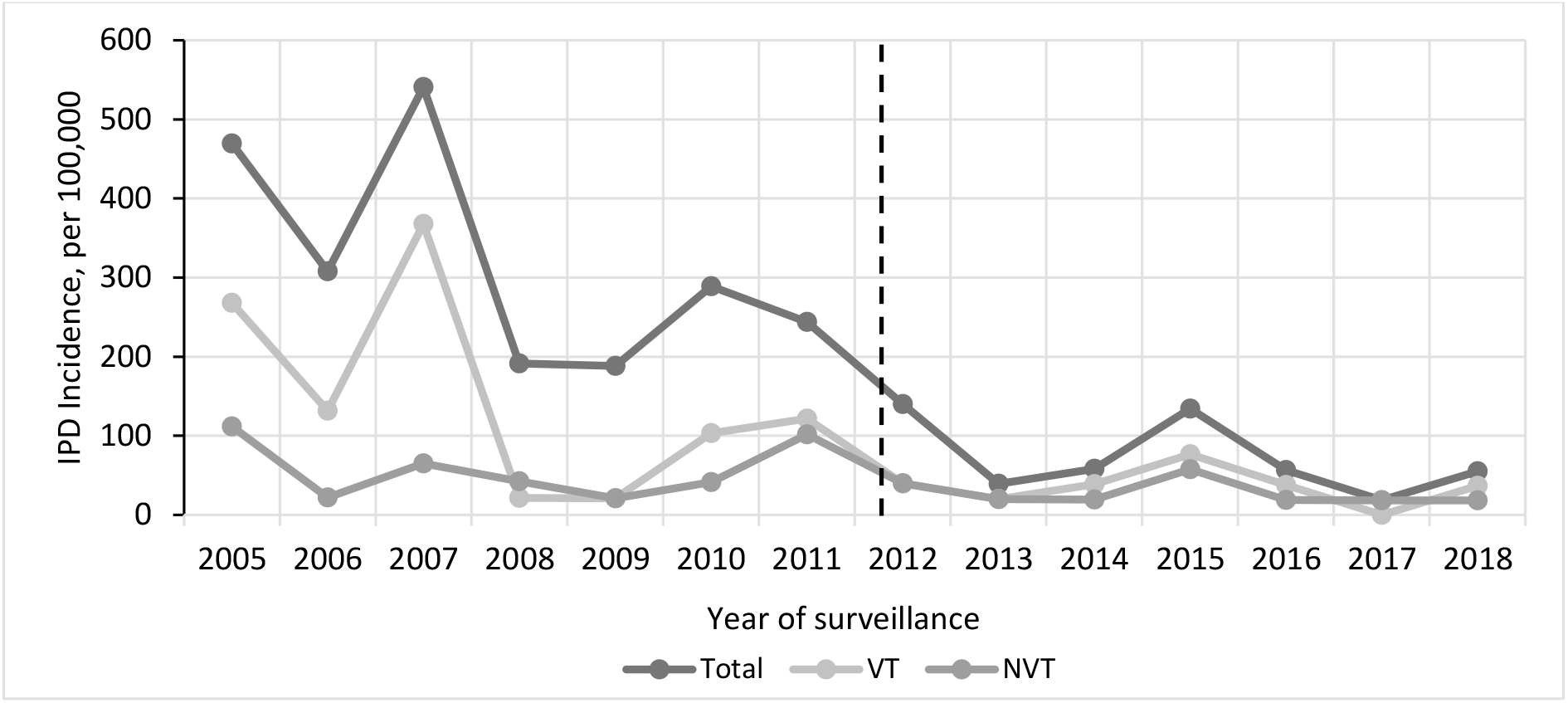
Incidence of invasive pneumococcal disease (IPD) in infants <90 days old (per 100,000 children <90 days old) in Blantyre, 2005 – 2018 Abbreviations: IPD, invasive pneumococcal disease; VT, vaccine type; NVT, non-vaccine type. Dashed line indicates date of PCV13 introduction. Refer to Supplementary Table S1 for incidence per year.

The mean incidence of total (VT+NVT) IPD cases in infants <90 days old in the pre-PCV13 period was 319 (95% CI 264-385) per 100,000 versus 72 (49-106) per 100,000 in the post-PCV13 period (P<0.01). For VT-IPD, the mean incidence pre-PCV13 was 148 (116-179) per 100,000 versus 36 (18-57) per 100,000 infants <90 days old in the post-PCV13 period. And for NVT-IPD the mean incidence pre-PCV13 was 58 (35-84) per 100,000 infants <90days old versus 27 (11-49) per 100,000 infants <90 days old in the post-PCV13 period. The overall reduction in total IPD incidence was 12% (IRR 0·88, 95% CI 0·86–0·90; p<0·001) per year among infants <90 days of age. For the post-PCV13 period there was a significant and further reduction of 46% (IRR 0.54, 0.46, 0.64; p<0.001).

## Discussion

In this low-income sub-Saharan African population with a high burden of disease, in which invasive pneumococcal disease incidence was already decreasing, we used our robust long-term hospital-based surveillance to show a substantial additional reduction in the incidence of vaccine serotype invasive pneumococcal disease among children aged <90 days following introduction of PCV13. However, our results also show that *Streptococcus pneumoniae* remains an important pathogen in causing bacteraemia and meningitis in neonates and young infants in Blantyre, Malawi.

The 7-valent pneumococcal conjugate vaccine was first introduced in the United States (US) in 2000 and resulted in a 45% overall reduction in VT-IPD (19). Several studies in other high-income settings have also reported significant reductions in IPD in neonates and young PCV-unvaccinated infants after introduction of PCV, suggesting protection through both direct and indirect effects. Ladhani et al (20) showed that in England and Wales introduction of PCV7 was responsible for an 83% reduction in VT-IPD in infants <90 days old and a declining trend in overall IPD. Similarly, studies in the US among neonates reported a 40-74% reduction in IPD after PCV7 introduction (21, 22).

Our study shows that in Blantyre, the incidence of IPD in young infants was already declining prior to PCV13 introduction on 12 November 2011, similar to the trend of decreasing IPD reported by Bar-Zeev *et al*. across all age groups in Blantyre (14). Similar findings have also previously been described in a longitudinal household study of pneumococcal acquisition among infants exposed to HIV in Karonga, northern Malawi (23) and in a study on paediatric bloodstream infections in Malawi (24). Possible explanations for this pre-PCV13 introduction secular decline in IPD are improved food security and nutrition in this population. Furthermore, from 2004 to 2015, the number of new patients started on ART in Malawi increased from about 3,000 to over 820,000, likely having an impact on the health of mothers and thereby improved infant wellbeing (25). Following the introduction of PCV13, IPD incidence in neonates and young infants in Blantyre further declined, consistent with an indirect effect of vaccination. However it is not possible to disassociate this from the pre-existing secular trend in this rare disease.

Nonetheless, VT-IPD remains present in this population, up to 7 years after PCV13 introduction. In our study more than 50% of cases in neonates and young infants in the post-PCV period were due to VT, with serotype 1 and 5 being most common both before and after PCV introduction. In sub-Saharan Africa, rates of pneumococcal carriage, which is a prerequisite for disease, are reported to be high with carriage rates of up to 90% described in the pre-PCV period (26-28). There is increasing evidence that in this setting, conjugate vaccines do not achieve the optimal reductions in VT pneumococcal carriage as seen in resource-rich countries. Recently, we have shown that in Malawi, despite high PCV13 uptake, there remains high persistent residual carriage of all PCV13 serotypes (12). Serotype 1, a common cause of IPD in Africa (29), was responsible for 3% of VT carriage of all ages in that study, consistent with our present findings in infants in this population. Furthermore another study in northern Malawi demonstrated a reduction in VT carriage after PCV13 introduction but found high carriage rates continued to be present among 6-week old infants too young to be vaccinated, with no difference in pneumococcal acquisition between the pre-and post-PCV13 period (28). The reduced vaccine impact on carriage in this setting is likely largely driven by demographic and socio-economic factors, and a local higher, age-dependent force of infection (30).

The majority (72%) of IPD cases in our study were of late onset disease, which is likely due to acquisition within households (31, 32). Previous studies in low income settings have described that pneumococcal acquisition occurs very early in life. Tigoi *et al* (33) showed that in Kilifi, Kenya the median time to acquisition was 38.5 days of life and Heinsbroek *et al* (23) showed a median time to first acquisition of 59 days of life in northern Malawi.

In our study, 28% of our cases occurred in the first 7 days of life with some occurring within the first 48 hours after birth, suggesting possible perinatal transmission during labour. Although *Streptococcus pneumoniae* is rarely isolated from the female genital tract, it is responsible for 1-11% of cases of neonatal sepsis (11, 34, 35). This indicates that mothers colonized with *Streptococcus pneumoniae* in the genital tract have a high likelihood of transmitting the organism to their infants at or very soon after birth and suggests a high invasion to colonization rate (2, 3, 36). Our results emphasize the need to prevent mother-to-child transmission and the importance of further research into preventative strategies such as maternal immunization.

Although this work provides a robust estimate of vaccine impact, it has several limitations. We were unable to review clinical, follow-up and outcome data as well as the individual vaccination status. Furthermore, our study only represents children who presented to hospital and not those managed in community health centres or at home, with a resulting risk of underestimation of true numbers. However, risk of underestimation is likely limited, given the clinical severity of IPD in infants and the fact that QECH is the only hospital in Blantyre District with inpatient paediatric facilities. Furthermore, there remains a risk of underestimation of IPD due to the challenges with blood volumes for culture in small infants. To our benefit, a large longitudinal study on bloodstream infections in children admitted to QECH has shown that the total paediatric and neonatal admissions have remained broadly constant since 2005 (24).

This study presents a unique set of data on the burden of IPD in infants less than 90 days old over a 14-year period. The results are derived from one of the few large databases across Africa based on long-term robust continuous prospective, systematic and reproducible surveillance. The Malawi-Liverpool-Wellcome Clinical Research Programme has provided routine, quality controlled, diagnostic blood culture service for paediatric patients with suspected severe bacterial infection admitted to QECH since 1998. Previously published robust surveillance studies at this site have shown similar methodology and consistency (15, 24).

## Conclusion

This study demonstrates that IPD incidence among neonates and young infants has declined over the past decade in Blantyre, Malawi. However, pneumococcal vaccine serotypes were the main cause of IPD both before and after PCV13 introduction. We postulate that there is incomplete indirect protection in this population, among whom many are too young to be vaccinated. Strategies such as maternal or neonatal immunization or schedule change with a booster dose to achieve greater reductions in the general population carriage need to be considered to protect this vulnerable population.

## Data Availability

The datasets generated during and/or analysed during the current study are available from the corresponding author on reasonable request.

## Ethical approval

The study proposal was approved by the College of Medicine Research and Ethics Committee, University of Malawi (P.02/15/1677) and the Liverpool School of Tropical Medicine Research Ethics Committee (14.056).

## Acknowledgments

We would like to thank the MLW laboratory management team (led by Brigitte Denis) and the MLW data management team (led by Clemens Masesa). Furthermore we would like to thank all the staff of the Chatinkha Neonatal Care Unit and the Paediatric Department at QECH for their efforts in caring for the most vulnerable infants and contributing to the collection of samples for this study.

## Supplementary Material

**Table S1.** IPD Incidence, per 100,000 population <90 days old, Blantyre Malawi

| Year | Total | VT | NVT |
| --- | --- | --- | --- |
| 2005 | 469.4 | 268.2 | 111.8 |
| 2006 | 307.8 | 131.9 | 22.0 |
| 2007 | 540.7 | 367.7 | 64.9 |
| 2008 | 191.6 | 21.3 | 42.6 |
| 2009 | 188.6 | 21.0 | 21.0 |
| 2010 | 288.8 | 103.1 | 41.3 |
| 2011 | 243.8 | 121.9 | 101.6 |
| 2012 | 140.1 | 40.0 | 40.0 |
| 2013 | 39.4 | 19.7 | 19.7 |
| 2014 | 58.3 | 38.9 | 19.4 |
| 2015 | 134.0 | 76.6 | 57.4 |
| 2016 | 56.6 | 37.8 | 18.9 |
| 2017 | 18.6 | 0.0 | 18.6 |
| 2018 | 55.1 | 36.7 | 18.4 |

